# Automated analysis of PSMA-PET/CT studies using convolutional neural networks

**DOI:** 10.1101/2021.03.03.21252818

**Authors:** Lars Edenbrandt, Pablo Borrelli, Johannes Ulén, Olof Enqvist, Elin Trägårdh

## Abstract

**Purpose:** Prostate-specific membrane antigen (PSMA) PET/CT has shown to be more sensitive and accurate than conventional imaging. Visual interpretation of the images causes both intra- and inter-reader disagreement and there is therefore a need for objective methods to analyze the images. The aim of this study was to develop an artificial intelligence (AI) tool for PSMA PET/CT and to evaluate the influence of the tool on inter-reader variability.

**Approach:** We have recently trained AI tools to automatically segment organs, detect tumors, and quantify volume and tracer uptake of tumors in PET/CT. The primary prostate gland tumor, bone metastases, and lymph nodes were analyzed in patients with prostate cancer. These studies were based on non-PSMA targeting PET tracers. In this study an AI tool for PSMA PET/CT was developed based on our previous AI tools. Letting three physicians analyze ten PSMA PET/CT studies first without support from the AI tool and at a second occasion with the support of the AI tool assessed the influence of the tool. A two-sided sign test was used to analyze the number of cases with increased and decreased variability with support of the AI tool.

**Results:** The range between the physicians in prostate tumor total lesion uptake (TLU) decreased for all ten patients with AI support (p=0.002) and decreased in bone metastases TLU for nine patients and increased in one patient (p=0.01). Regarding the number of detected lymph nodes the physicians agreed in on average 72% of the lesions without AI support and this number decreased to 65% with AI support.

**Conclusions:** Physicians supported by an AI tool for automated analysis of PSMA-PET/CT studies showed significantly less inter-reader variability in the quantification of primary prostate tumors and bone metastases than when performing a completely manual analysis. A similar effect was not found for lymph node lesions. The tool may facilitate comparisons of studies from different centers, pooling data within multicenter trials and performing meta-analysis. We invite researchers to apply and evaluate our AI tool for their PSMA PET/CT studies. The AI tool is therefore available upon reasonable request for research purposes at www.recomia.org.

## 1. Introduction

Prostate-specific membrane antigen (PSMA)-ligand positron emission tomography/computed tomography (PET/CT) imaging has shown to be of value for men with prostate cancer both for primary staging and in patients with biochemical recurrence. PSMA PET/CT has shown to be more sensitive and accurate than the conventional imaging standard of CT and bone scan^1-8^. It is also more cost-effective compared with conventional imaging^9^.

The interpretations of PSMA PET/CT images, both clinically and in research, are based on visual analysis. Lesions are detected by an image reader and in cases quantification of disease burden is done, easily accessible measurements such as the maximum standardized uptake value (SUV) are usually used. Standards for reporting have been proposed in order to facilitate comparison of results from different studies^10-12^. Despite these efforts, visual interpretation of PSMA PET/CT has shown to cause both intra- and inter-reader disagreement even in single site studies^13,14^. The disagreement is most likely worse if readers from different hospitals are compared. There is an unmet need for objective methods to analyze PSMA PET/CT studies to improve reproducibility. Artificial intelligence (AI) based methods has the potential to fill this clinical need^15,16^. An AI-based method can also calculate measurements reflecting the entire disease burden in the body. Volume and tracer uptake in automatically detected tumor lesions can be calculated, a task too time-consuming to perform manually in clinical practice. Total lesion uptake (TLU) most likely reflects tumor burden more accurately than maximum SUV, which is based on a limited volume of the tumor^17^.

We have recently showed that AI tools based on convolutional neural networks can be trained to automatically segment organs, detect tumors, and quantify volume and tracer uptake of tumor lesions in PET/CT. In one study AI-based measurements of bone metastases showed to be associated with overall survival^18^. In a second study AI-based quantification of the primary tumor in the prostate gland showed to be associated with overall survival^19^. A third study showed that AI-based detections of lymph nodes were associated with prostate cancer specific survival^20^. These studies were based on the non-PSMA-targeting PET tracers choline and sodium fluoride for imaging prostate cancer. The retrospective design of those studies gave us the opportunity to evaluate the AI tools using survival data and not only comparisons with human experts. The disadvantage being that PSMA PET/CT has emerged the most promising PET tracer for imaging prostate cancer patients.

The aim of this study was to develop an AI tool for PSMA PET/CT based on our previous AI tools and to evaluate the influence of the AI tool on inter-reader variability for quantification of primary prostate tumors and bone metastases, and detection of lymph nodes.

## 2. Methods

### 2.1 Patients and Imaging

The AI tool was applied to ten patients with high-risk prostate cancer referred for PSMA PET/CT for initial staging at Skåne University Hospital, Lund and Malmö, Sweden during 2019. Patients were administered with 4 MBq/kg [18F]-PSMA-1007 and after 120 min scanned on a Discovery MI PET/CT (GE healthcare, Milwaukee, WI). The patients were scanned from the mid-thigh to the base of the skull. Images were acquired for 2 min/bed position. The PET images were reconstructed using the commercially available block-sequential regularization expectation maximization algorithm Q.Clear (GE Healthcare, Milwaukee, WI) including the time-of-flight and point spread function modelling with a 256 x 256 matrix (pixel size 2.7 x 2.7 mm2, slice thickness 2.8 mm) and a beta factor of 800^21^.

The study was conducted according to the principles expressed in the Declaration of Helsinki, and approved by the local research ethics committee at Lund University (#2016/193 and #2018/753). All patients provided written informed consent.

### 2.2 AI Tool

The new AI tool relies heavily on the CT-based organ segmentation tool from work by Trägårdh et al. ^22^. This tool is used to find all relevant bones, the prostate, the urinary bladder and other relevant organs.

#### 2.2.1 Prostate tumor

The lesion threshold is set to SUV of the first quartile of the prostate uptake + 2, but at most SUV=5. Any high uptake region connected to the prostate is included unless it originates from bone, bladder or gastrointestinal tract.

#### 2.2.2 Bone metastases

The normal range of SUV varies throughout the bones in the body. To handle this, the bones are organized into the following groups: vertebra, rib, scapula, radius, fibula, patella, hand, humerus, clavicle, tibia, foot, hip, femur, ulna and sternum. For each of the groups, the lesion threshold is set to SUV of the median uptake * 3, restrained between SUV interval of 2-10.

#### 2.2.3 Lymph nodes

It is very difficult to segment lymph nodes directly in CT images. Instead, we trained the AI tool to directly segment lymph nodes in 18F-fluorocholine PET/CT using an organ mask, the CT image and the PET image as input as described in the work by Borrelli et al. ^20^.

#### 2.2.4 Postprocessing

Both the methods for prostate tumors and bone metastases rely directly on a CT segmentation to define the area of interest, making them sensitive to PET/CT misalignment. To reduce this problem, each pixel is assigned to a close local maximum using a flooding algorithm. Uptake in a pixel is assumed to originate from the organ of the corresponding local maximum.

As the final step for all three methods, lesions with TLU less than 2.0 were removed.

### 2.3 Manual analysis

Three nuclear medicine specialists from two different hospitals independently segmented primary prostate tumors, bone metastases, and lymph nodes in the ten PSMA PET/CT studies, first without AI support and then, more than eight weeks later, with segmentations proposed by the AI tool. The physicians were informed that TLU measurements were to be calculated based on their segmentations of prostate tumors and bone metastases. For lymph nodes no TLU measurements were to be calculated and they were only instructed to mark detections nodes. The cloud-based annotation platform RECOMIA (https://www.recomia.org) was used for the manual segmentations. The platform includes basic display features for PET/CT images and segmentation tools (Fig 1). AI segmentations can be viewed as an overlay on PET and CT images and in 3D images. The AI detections could easily be deleted or adjusted by the physicians and additional lesions could be segmented.

**Fig 1.**
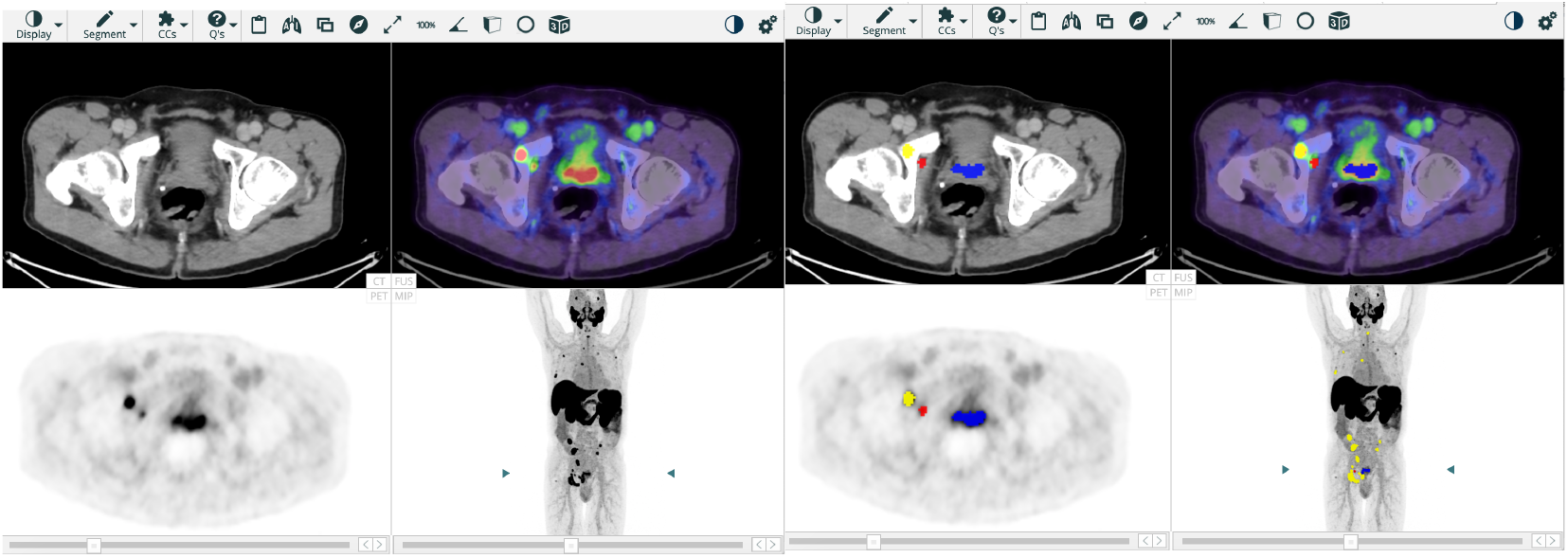
PET/CT display without (left) and with (right) segmentations of prostate tumor (blue), bone metastases (yellow), and lymph nodes (red).

### 2.4 Statistical Analysis

To measure the inter-reader variability in TLU measurements for each patient, the range of the TLU measurements from the three observers was computed. A two-sided sign test was used to analyze the number of cases with increased and decreased variability with support of the AI tool.

For lymph nodes the number of detections was analyzed instead of TLU measurements as that number has shown to be associated with survival in our previous work^20^. If an AI detected lymph node has non-zero overlap with a manually detected lymph node it is considered a true positive and otherwise a false negative lymph node. AI detected lymph nodes with no overlap with a manually detected lymph node are considered false positives. As there are three manual segmentations without AI support, we compare the AI to one manual reference at a time and then average the results over the three choices of reference. Inter-observer variability is measured analogously. Each possible reference is compared to the other two manual segmentations and the pairwise results are averaged. The effect of having AI support was measured in a similar way. One segmentation is held as reference and results are averaged over the six different references.

## 3. Results

### 3.1 Prostate tumors

The AI-based prostate tumor TLU measurements are compared to the corresponding fully manual measurements from the three physicians in Figure 2. In all ten patients, the AI tool gives a value within the range of the three physicians.

**Fig 2.**
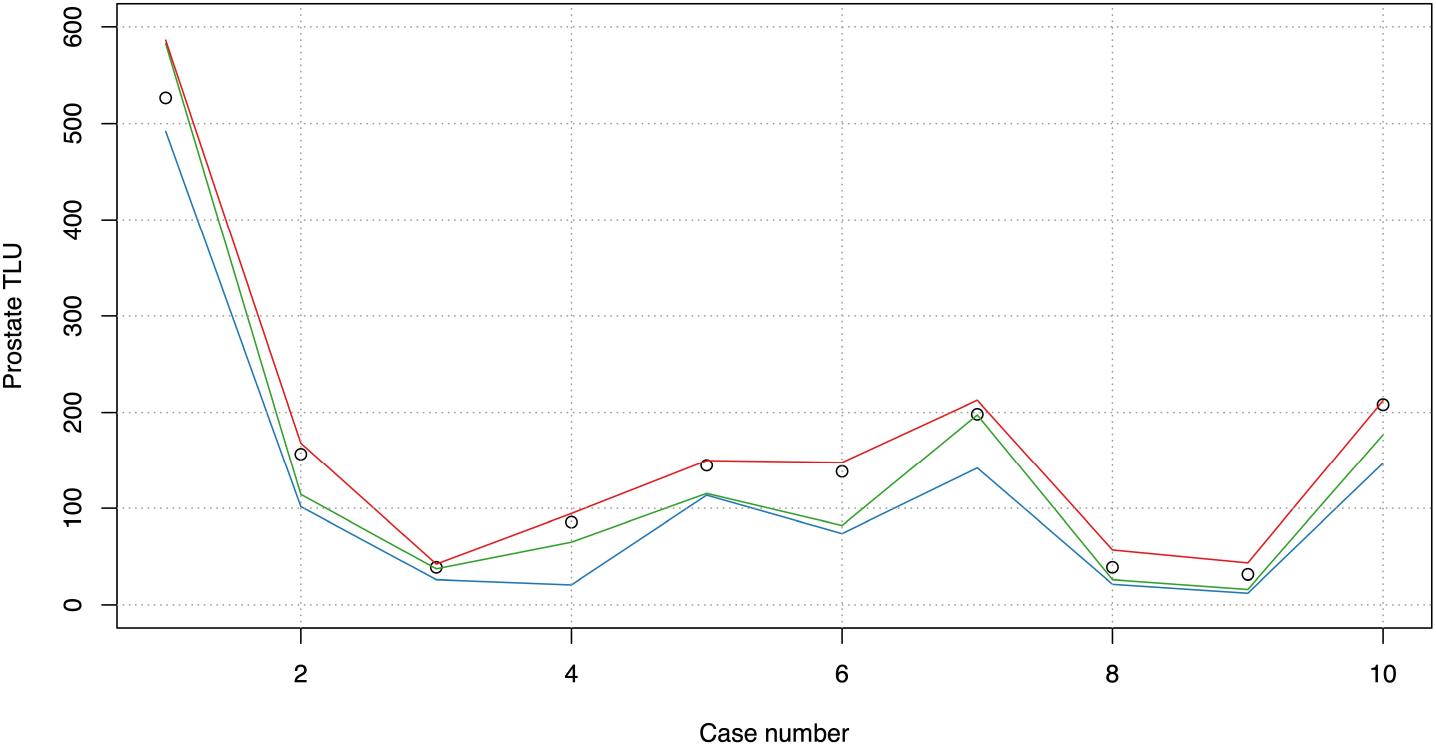
Prostate tumor TLU values for the AI tool (black rings) compared to the minimum (blue line), median (green line) and maximum (red line) values from the three physicians without AI support.

The three physicians produced more consistent prostate tumor TLU measurements with AI support than without AI support for all ten patients (p=0.002), see Fig. 3.

**Fig 3.**
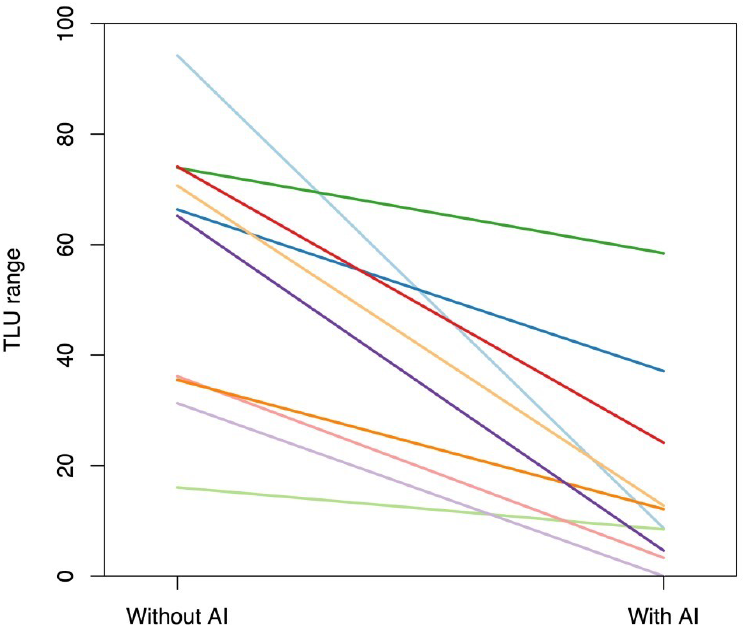
The range of prostate tumor TLU measurements by the three physicians in ten patients without AI support to the left and with AI support to the right.

### 3.2 Bone metastases

The AI-based bone metastases TLU measurements are compared to the corresponding fully manual measurements from the three physicians in Figure 4. In eight of ten patients, the AI tool gives a value within the range of the physicians and in two cases it gives a slightly larger value.

**Fig 4.**
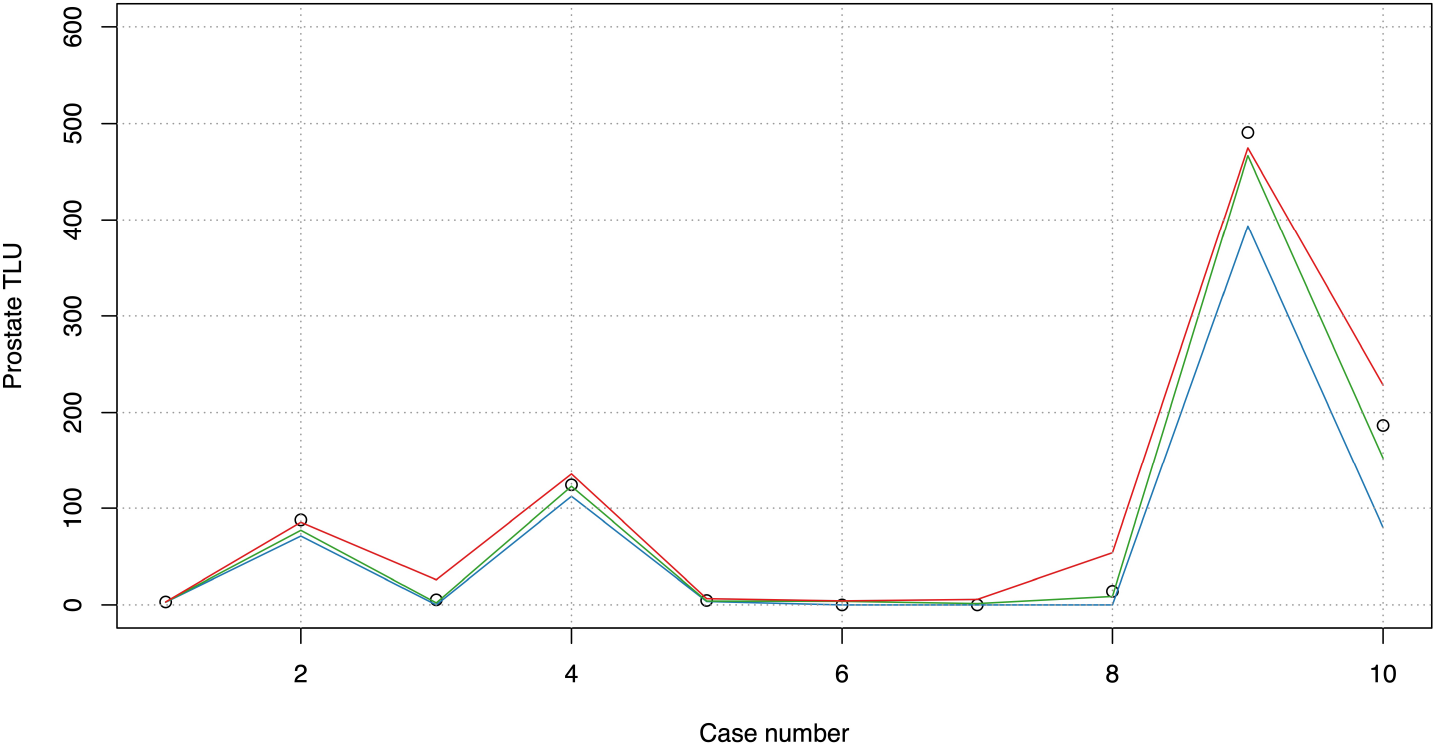
Bone metastasis TLU values for the AI (black rings) compared to the minimum (blue line), median (green line) and maximum (red line) values from the three physicians without AI support.

AI support decreased the range in bone metastasis TLU measurements in nine patients and increased the range in one patient (p=0.01), see Fig. 5.

**Fig 5.**
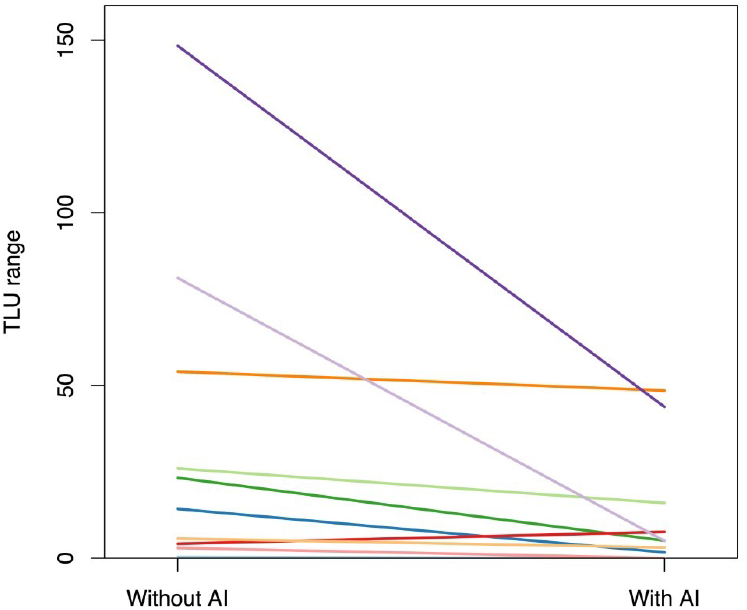
The range of bone metastases TLU measured by the physicians in ten patients without AI support to the left and with AI support to the right.

### 3.3 Lymph nodes

On average a reader detected a total of 74 (66-80) lymph nodes in the ten patients without AI support. The AI tool detected on average 48 (43-56) of these giving an average sensitivity of 66% (54%-74%). Apart from that the AI tool detected an average of 26 (20-37) false positive lesions, i.e., 2.6 per patient. The average positive predictive value was 63% (59%-66%). As a comparison the average inter-observer sensitivity was 80% (59%-95%) and the average inter-observer positive predictive value was 81% (65%-92%).

Without AI support, all three physicians agreed in on average 72% of the lymph nodes. With AI support this number decreased to 65%. In the remaining cases two physicians were in agreement, and one reader had a different opinion. Note that physicians can either agree that an uptake is not a lymph node or agree that it is.

## 4. Discussion and Conclusions

Physicians supported by an AI tool for automated analysis of PSMA-PET/CT studies showed significantly less inter-reader variability in the quantification of primary prostate tumors and bone metastases than when performing a completely manual analysis. A similar effect was not found for lymph node lesions. The range in TLU measurements for prostate tumors and bone metastases and the agreement on only 72% of the lymph nodes between the three physicians without AI support illustrate the problem with inter-reader disagreement, a problem that has been described in previous studies^13,14^.

An AI tool has also shown to be more reproducible than physicians in the task to segment bones in two different CT from the same patient^23^. This suggests that there is a need for automated, objective, and reproducible image analysis. AI tools offer an interesting opportunity to improve reproducibility and reduce reading times for PET/CT analysis.

A high false positive rate of detections has been a major limitation for clinical AI tools in mammography and lung imaging. It can be time-consuming for radiologists to review and dismiss large number of false positive detections and there is also a risk that it leads to decreased specificity^24^. The low number of false AI detections of lymph nodes in this study, 2.6 per patient, was therefore encouraging.

Limitations of this study include the small number of PMSA PET/CT studies and physicians. It is also based on one of several available PSMA tracers. The performance of the AI tool in PSMA PET/CT studies based on other PSMA tracers need to be evaluated.

We have used non-PSMA-targeting tracers to train and evaluate our first AI tools for automated analysis of PET/CT studies from patients with prostate cancer. This gave us the opportunity to assess the clinical value of the tools using OS or prostate cancer specific survival as reference. PSMA PET/CT has now been used for relatively long time that it will be possible to assess the clinical value of AI tools for PSMA PET/CT.

It is interesting that our AI tool developed for non-PSMA-targeting tracers showed such a good performance when applied to PSMA PET/CT studies. Some features of abnormal lymph nodes such as size, shape, and localization are the same in PET/CT studies based on PSMA and non-PSMA-targeting tracers. The tracer distribution is, however, different and the performance will most likely improve when PSMA PET/CT studies are used to train the AI tool. We have therefore trained a new version of the AI tool with close to 200 PSMA PET/CT studies with manual segmentations. This version 1.1 is now available for research at our platform.

In conclusion, we have presented an AI tool for automated analysis of PSMA PET/CT, which decreased inter-reader variability in the quantification of primary prostate tumors and bone metastases. The tool may facilitate comparisons of studies from different centers, pooling data within multicenter trials and performing meta-analysis. We invite researchers to apply and evaluate our AI tool for their PSMA PET/CT studies. The AI tool is therefore available upon reasonable request for research purposes at www.recomia.org.

## Data Availability

The AI tool and data presented in this study is available upon reasonable request for research purposes at www.recomia.org.

